# The Metabolic Role of MAP3K15: Genetic and Phenotypic Insights from the 23andMe Research Database and Genetics-Driven Recruitment

**DOI:** 10.1101/2024.01.09.24301012

**Authors:** Jennifer J. Brady, Kira Kalkus, Dominique T. Nguyen, Jingran Wen, Zachary L. Fuller, Yue Qi, Shaeloren S. Deering, Leah Selcer, Suyash S. Shringarpure, The 23andMe Research Team, Michael V. Holmes, Xin Wang

## Abstract

MAP3K15 has been previously associated with protection from type 2 diabetes (T2D), prompting interest in the development of MAP3K15 inhibitors as a potential therapeutic option for diabetes. The trans-ancestry genome-wide association study (GWAS) meta-analysis and loss-of-function (LoF) burden testing methods that implicate association with T2D greatly benefit from large sample size. The direct-to-consumer genetic testing company, 23andMe, Inc., is the world’s largest research consented genetic database. We leveraged the 23andMe database to further inform the metabolic role of MAP3K15, using a variety of genetic analysis methods. We find that MAP3K15 LoF carriers show a significant delay of 4.5 years in the median age of T2D diagnosis among individuals at high polygenic risk and uncover a novel burden association of MAP3K15 LoF with protection against high cholesterol. We expanded these findings by establishing a capability to recruit consented participants on the basis of genetics unknown to them (specifically, a single LoF variant in MAP3K15, rs148312150), and obtained clinical laboratory evidence of a modest reduction in median cholesterol and LDL/HDL ratio in MAP3K15 LoF carriers. Our findings demonstrate the discovery power of the 23andMe database, including the feasibility of consented participant recruitment to inform therapeutic discovery and development.

## Introduction

The 23andMe database is a vast and dynamically updated repository of deeply genotyped and phenotyped individuals. At the time of writing, the 23andMe database had over 11 million customers consenting to participate in research, with 5.2 million individuals consenting to be recontacted on the basis of genetics unknown to them (i.e., genetics that are not disclosed in 23andMe consumer product offerings).

Characterization of unique genetic variants plays a key role in the success of novel therapeutic discovery and development. Deep phenotyping of individuals with natural mutations of interest (such as human genetic knockouts) can suggest new disease therapeutic areas and help inform the efficacy and potential toxicity of a new drug prior to significant pipeline investment^1,2^. The benefits of deep phenotyping also extend to late-stage drug development, such as enabling the modeling of control and treatment arms to improve clinical trial design, the identification of exploratory biomarkers, or selection of patients for clinical trials^3^.

In drug discovery, the phenotypes of individuals with LoF variants can provide information on safety risks as well as identify indication expansion opportunities for a target therapeutic. A notable example of how phenotyping of a single LoF individual enabled the pharmaceutical industry to de-risk clinical development came from the identification of a very rare compound heterozygous female for PCSK9 LoF in the Dallas Heart Study^4^. PCSK9 gain-of-function variants were known to be causal for hypercholesterolemia^5^, suggesting that PCSK9 inhibition could be of therapeutic value. However, the broad tissue expression of PCSK9, including brain, kidney, and reproductive organs gave the pharmaceutical industry pause due to the unknown potential for on-target toxicity. One such concern was the risk that PCSK9 inhibition might lead to neurocognitive decline^6^. The discovery of a PCSK9 knockout female in good overall health with normal kidney and liver function, a college education, and two young children alleviated the major safety-risk concerns, resulting in the development of several therapeutic programs, FDA approvals, and ongoing clinical trials targeting PCSK9 with various modalities^1,7^.

LoF variants are typically rare (e.g., frequency ≤ 0.01%)^8,9,10^ and it can be difficult to have sufficient statistical power to detect associations with phenotypes of interest at individual variants. Gene-based burden tests are a popular alternative approach, where evidence is aggregated across variants of a given class (e.g., LoFs) in a gene, thereby boosting the statistical power to detect associations^11,12^. The size of the 23andMe database and imputation panel can be leveraged to identify rare knockout individuals. In particular, X chromosome LoF variants present a unique situation of hemizygosity: males with a single protein-truncating variant have a complete loss of function, enabling inference on the effect of complete gene inactivation more readily than in females or from autosomal genes, where homozygous carriers tend to be too infrequent^13,14^.

While rare variants, such as LoFs, can have large effects on complex traits, they typically only explain a small amount of population variance^15,16,17^. On the other hand, common variants tend to have small effect sizes individually on a given phenotype, yet in aggregate explain a large proportion of the observed heritability. Polygenic risk scores (PRS) derived from these common variants have proven to be powerful predictors of disease risk and can stratify patients into clinically relevant groups^18^. Recent work has demonstrated that rare penetrant mutations combined with a common variant PRS are best powered to predict disease status and to identify individuals at the extreme tails of the phenotypic distribution^19^. Moreover, large effect rare variants have been shown to modify disease risk for individuals with extremely high or low PRS^20^.

Here we leverage the 23andMe Research Cohort to derive insights into the metabolic role of the X chromosome gene MAP3K15, both within the 23andMe database and by deep phenotyping of knockout individuals through genetics-driven recruitment.

## Methods

### Study participant selection and ethics approvals

We recruited rs148312150 LoF carriers and age- and sex-matched controls from the consented 23andMe Research Cohort. We restricted our re-contacting of individuals on the basis of their geographic location (self-reported zip code) to ensure they were within a fifty mile radius of an area with mobile phlebotomy-coverage. Prospective participants were recruited via email and directed to the study landing page for additional study information and to complete eligibility screening. Eligible individuals included those who: lived in the United States, were at least 18 years of age, consented to 23andMe Research, consented to the 23andMe Opt-In for Recruitment Based on Unreported Genetics, could consent for themselves, were willing to provide a blood sample for genetics analyses, clinical lab values, and other biological analyses, were able to read and write English fluently, had internet access, and had not had a vasovagal response from any previous blood draw procedure. Participants provided informed consent and volunteered to participate in the research online under a protocol approved by the external AAHRPP-accredited Salus IRB (https://www.versiticlinicaltrials.org/salusirb). All research was performed in accordance with relevant guidelines/regulations, including the Declaration of Helsinki.

### Sequencing validation

Genomic DNA was purified from fresh whole blood samples sent to 23andMe using the DNeasy Blood and Tissue Kit (Qiagen). Purified gDNA was subject to PCR amplification using primers flanking rs148312150. Variant status (control versus knockout) was confirmed by Sanger Sequencing, with 100% concordance between the 23andMe assigned genotype and sequencing results (data not shown).

### Statistical analyses

Data were analyzed using Python (version 2.7.18), with figures generated using matplotlib (version 2.0.2). Genetically inferred ancestry was obtained from the 23andMe database^21^. Data are presented as median, interquartiles, and range. Comparisons between control and knockout groups were conducted using the Mann-Whitney U test. Correction for multiple testing was not performed due to the strong prior from burden tests and the highly correlated nature of the lipid species analyzed.

### Burden testing

The association between imputed rare variants and phenotypes was tested on a per-gene basis using a burden test. We first annotated variants with VEP to obtain protein consequences according to the Ensembl v.109 canonical transcript. Additionally, LOFTEE v.1.0.4^22^ was used to annotate predicted loss-of-function variants (pLoFs) as “high-confidence” or “low-confidence” under default settings. For inclusion in the burden test, we considered variants with a minor allele frequency (MAF) < 1% and annotated as “high-confidence” (HC) LoF. To calculate a burden score for each gene, we apply the common collapsing approach^23^, assuming that each variant has the same magnitude and direction of effect. Such a collapsing approach is typically applied for burden tests using genotyped variants. Here, we apply a probabilistic version of this collapsing method for use with imputed haploid dosages. We calculate the probability of each haplotype containing at least one qualifying variant and take the sum from both haplotypes as the burden score. Specifically, for individual sample *i*, the burden score *S* is calculated as

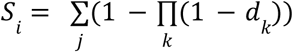

for each haplotype *j* where *d* represents the imputed dosage of each qualifying variant *k* in the gene.

Case-control phenotypes used in the association analyses are in a European cohort, controlling for age, sex and five genetic principal components in a penalized logistic model. The burden association is adjusted for nearby common variant associations by including effectively independent genome-wide association study (GWAS) hits identified by conditional fine-mapping^24^ on the same chromosome that have *p* < 1e-7 as covariates, following the approach of Backman *et al.* (2021)^25^.

### Polygenic risk score

We constructed a polygenic risk score for type 2 diabetes diagnosis using weights available from the publicly available PGS Catalog^26^ under accession ID PGS000713. Specifically, the weights available from the PGS Catalog were originally estimated by Sinnot-Armstrong et al., (2021)^27^ for 183,830 directly genotyped variants from 223,327 individuals of British ancestry available from the UK Biobank. No MAP3K15 variants were included in the PRS, as only autosomal variants were included in the original study.

## Results

We examined phenotypic associations with MAP3K15 in the 23andMe database and observed replication of a previous association with T2D from a trans-ancestry genome-wide meta-analysis^28^, in addition to observing other diabetes-related phenotypes, such as gestational diabetes (**Figure 1**). We found a clear genome-wide significant single-variant association of MAP3K15 with T2D. Of note, the T2D association includes the MAP3K15 coding variant rs148312150 (p.Arg1136Ter), a rare (MAF = 0.453%) stop-gain LoF variant (predicted based on sequence context) with a protective odds ratio (odds ratio = 0.824, 95% CI: [0.787,0864], p = 2.7e-17). T2D often presents with comorbidities including obesity, hypertension, dyslipidemia, and heart disease. The association of MAP3K15 with hypertension was observed in the opposite direction compared to diabetes phenotypes (odds ratio = 1.049, 95% CI: [1.036,1.061], p = 2.6e-14), consistent with previous observation of elevated systolic blood pressure of sodium challenged MAP3K15 knockout mice^29,30^. LoF burden test (odds ratio = 1.031, 95% CI: [1.004,1.017], p = 0.0101) shows consistent directionality with the single variant association. This contrasts the sub-significant associations with hypertension (odds ratio = 0.90, 95% CI: [0.80,1.02], p = 0.10) and the modest effect size estimate for systolic blood pressure (beta = −0.07, 95% CI: [−0.12,-0.01], p = 0.01) reported in MAP3K15-LoF burden analyses of hemizygous male knockout individuals in the UK Biobank^30^.

**Figure 1.**
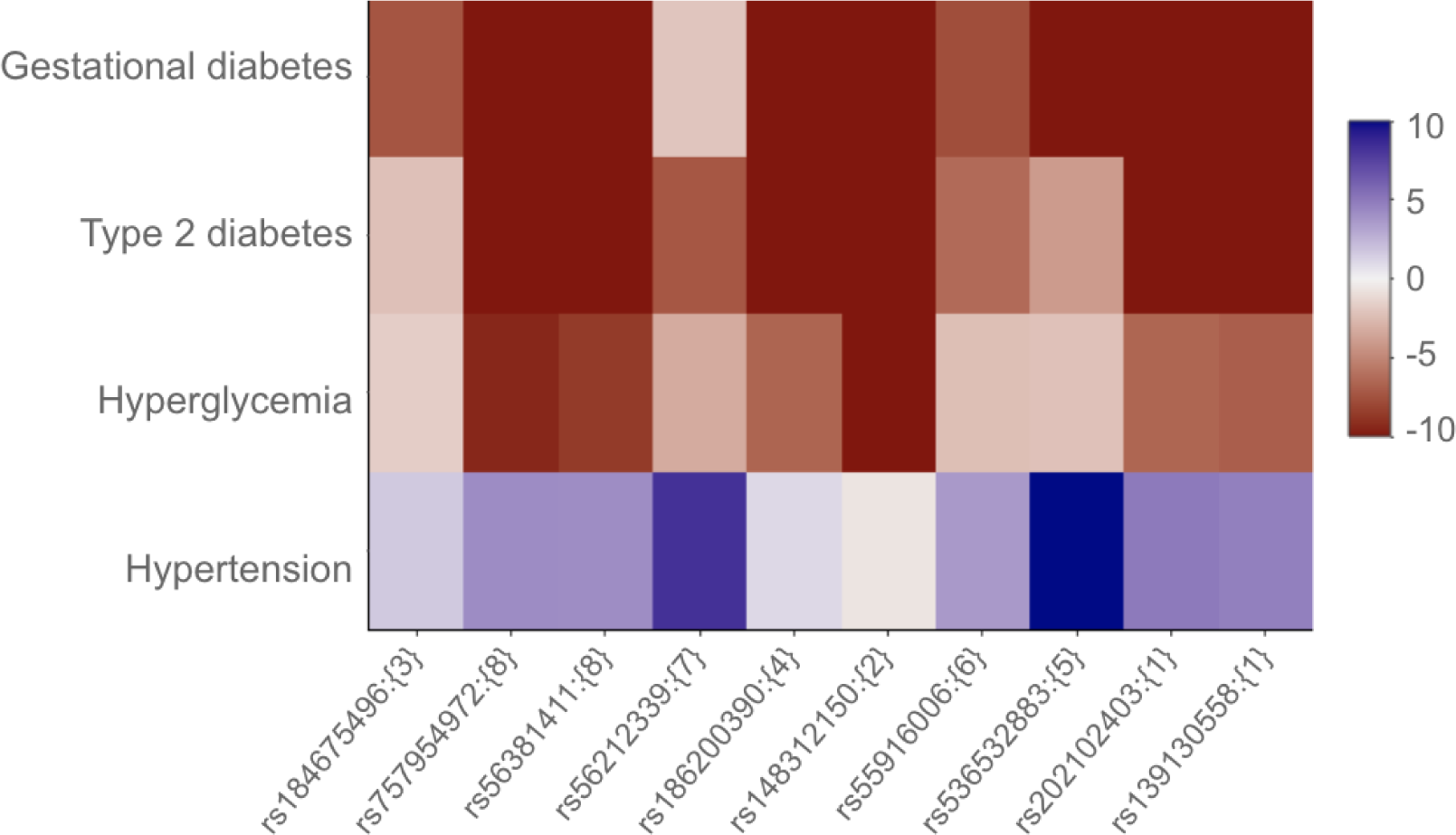
Metabolic phenotypes associated with MAP3K15 in the 23andMe database. The heatmap shows significance and direction of the associations with MAP3K15. The blue-red coloring corresponds to the direction of effect that enforces consistency among highly correlated SNPs. The intensity of the colors correspond to -log_10_(p-value) that is capped at 10. SNPs are ordered based on LD, and clustered into groups for representation (curly brackets identify cluster IDs). Only associations with p-value < 5e-8 are included.

Whole exome sequencing and LoF burden testing in the UK Biobank and FinnGen datasets have previously identified associations of MAP3K15 LoF with protection from T2D and elevated hemoglobin A1c (HbA1c)^25,30^. We replicated the protective associations for T2D and related glucose phenotypes with MAP3K15 LoF variants via burden testing in the 23andMe database (**Figure 2** and **Extended Data Table 1**). The association of MAP3K15 LoF with protection with T2D persists after adjusting for BMI^31^ and these data are replicated at 23andMe (odds ratio adjusted for BMI = 0.824, 95% CI: [0.803,0.844], p = 1.85e-50). Moreover, the burden association remains significant (odds ratio = 0.847, 95% CI: [0.826-0.869], p = 1.81e-34) even if the stop-gain LoF implicated in the GWAS is removed from the set of variants, providing further evidence that loss-of-function of MAP3K15 is associated with protection against T2D. Together, these results highlight the strength of the large sample size available in the 23andMe database, where rare variants tested individually with GWAS and in aggregate with burden tests can reveal associations with human disease.

**Figure 2.**
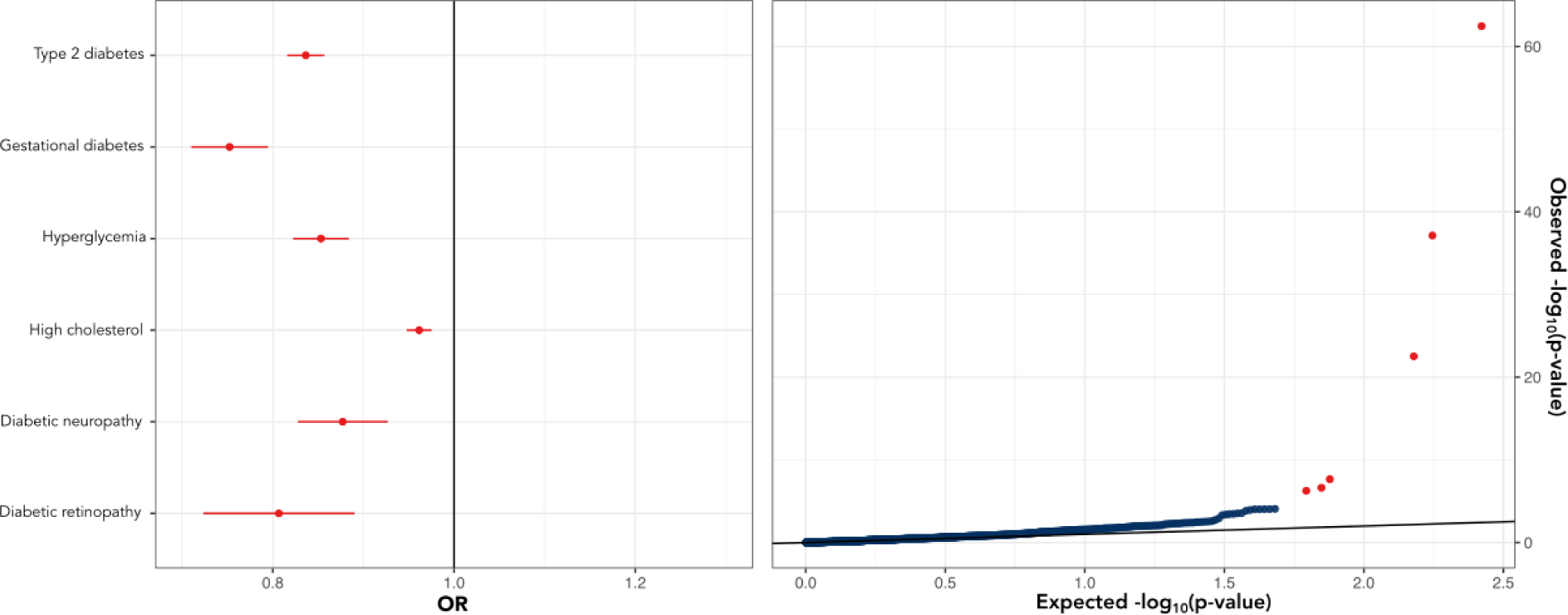
MAP3K15 loss-of-function is associated with protective effects for T2D-related phenotypes and high cholesterol. Forest plot of MAP3K15 LoF associations, odds ratios and 95% confidence intervals are shown (left). Quantile-quantile plot (Q-Q) plot for MAP3K15 loss-of-function burden association for 1,149 binary case-control phenotypes (right). The red dots represent the six significantly (p < 1e-7) associated phenotypes in the same order listed on the left, while the black line is the expectation under the null hypothesis of uniformly distributed p-values.

Intriguingly, we also uncovered a novel statistically significant protective association for high cholesterol (odds ratio = 0.961, 95% CI: [0.948,0.975], p = 2.2e-8, **Extended Data Table 1**) that was not found in previous studies using FinnGen and UK Biobank. Although the MAP3K15 LoF association with cholesterol from UK Biobank is far from significant (p-value burden = 1.24e-1, https://app.genebass.org), it is worth noting that the association is directionally consistent with a protective association for cholesterol (beta burden = −7.4e-4)^32^. We suspected that our ability to discover this association was due to the unique power from the 23andMe data. We hence conducted a power analysis. In the UK Biobank, there are multiple measures of cholesterol, including self-reported cholesterol diagnosis and quantitative measurements of cholesterol from blood biochemistry laboratory values. The 23andMe phenotype takes into account both high cholesterol diagnosis and treatment, defining cases as those who report ever having a high cholesterol diagnosis or having been on any cholesterol-lowering medication, with controls reporting neither. Our analysis compares the self-reported high cholesterol phenotype from 23andMe with self-reported “high cholesterol” from the UK Biobank (Field: 20002_1473). Using an odds ratio of 0.961 (as estimated from 23andMe data), we quantified a power (alpha = 1e-7) of 0.88 to find a significant association in 23andMe, compared to 2e-5 in the UK Biobank.

We set out to investigate in more detail which factors might have led to the differential power in the two datasets. The main driver for the differential discovery power is the larger cohort size and prevalence of individuals with high cholesterol in the 23andMe database. The 23andMe high cholesterol phenotype has a total sample size of 2,844,426, with a prevalence of ∼44%. From UK Biobank data, the sample size for the high cholesterol phenotype is an order of magnitude less, with only 394,783 individuals and a prevalence of ∼12%. The difference in prevalence of self-reported high cholesterol between 23andMe and the UK Biobank may be attributable to different possible definitions of high cholesterol and/or treatment decisions in relation to lipid-lowering therapies among research participants. For example, the 23andMe phenotype includes individuals on medication for high cholesterol as cases. Another important factor for the difference in discovery power is the greater cumulative allele frequency observed in the 23andMe cohort. The observed cumulative allele frequency for MAP3K15 LoF variants in the UK Biobank was ∼0.6%, while the total cumulative imputed allele frequency was 1.1% in the 23andMe data. As such, the UK Biobank is severely underpowered to detect an association between MAP3K15 LoF and high cholesterol. To further take into account these differing factors, we performed calculation adjustments to the UK Biobank data to match the 23andMe prevalence and cumulative allele frequency (while keeping the total sample size the same as 23andMe). Even with these adjustments the power to detect an odds ratio of 0.961 remains low (0.02). In short, it was not possible to make this discovery using the comparable self-reported UK Biobank data.

In order to gain additional insight into the nature of how MAP3K15 LoF offers protection against diabetes, we examined the effect of MAP3K15 LoF in approximately 3.6 million samples from the 23andMe database predicted to be at high or low risk of T2D diagnosis on the basis of their common polygenic risk quantified by a T2D PRS. Specifically, we removed the MAP3K15 locus from the T2D PRS and compared MAP3K15 LoF carriers and non-carriers in the 5th and 95th percentile of the T2D PRS distribution. For each PRS group, we then modeled the proportion of individuals diagnosed with T2D as a function of their self-reported age of diagnosis. Across all T2D cases, the median age of diagnosis was 50. As expected, the age of T2D onset for individuals at low risk for T2D (i.e., in the bottom 5% of the PRS distribution) did not show strong dependence on MAP3K15 LoF, with no significant effect of MAP3K15 LoF carrier status on the proportion diagnosed (p = 0.32, log-rank test). However, for individuals at high risk for T2D (i.e., in the top 5% of the PRS distribution), stratification by MAP3K15 LoF status had a significant effect on the proportion of T2D cases (p = 0.029, log-rank test), with LoF carriers having a significantly delayed age of diagnosis (Event Time Ratio [95% CI]: 1.27 [1.02-1.58]). Among those with high polygenic risk, MAP3K15 LoF non-carriers and carriers have a median age of T2D diagnosis of 46 and 50.5, respectively, representing an effective 4.5-year delay (**Figure 3**).

**Figure 3.**
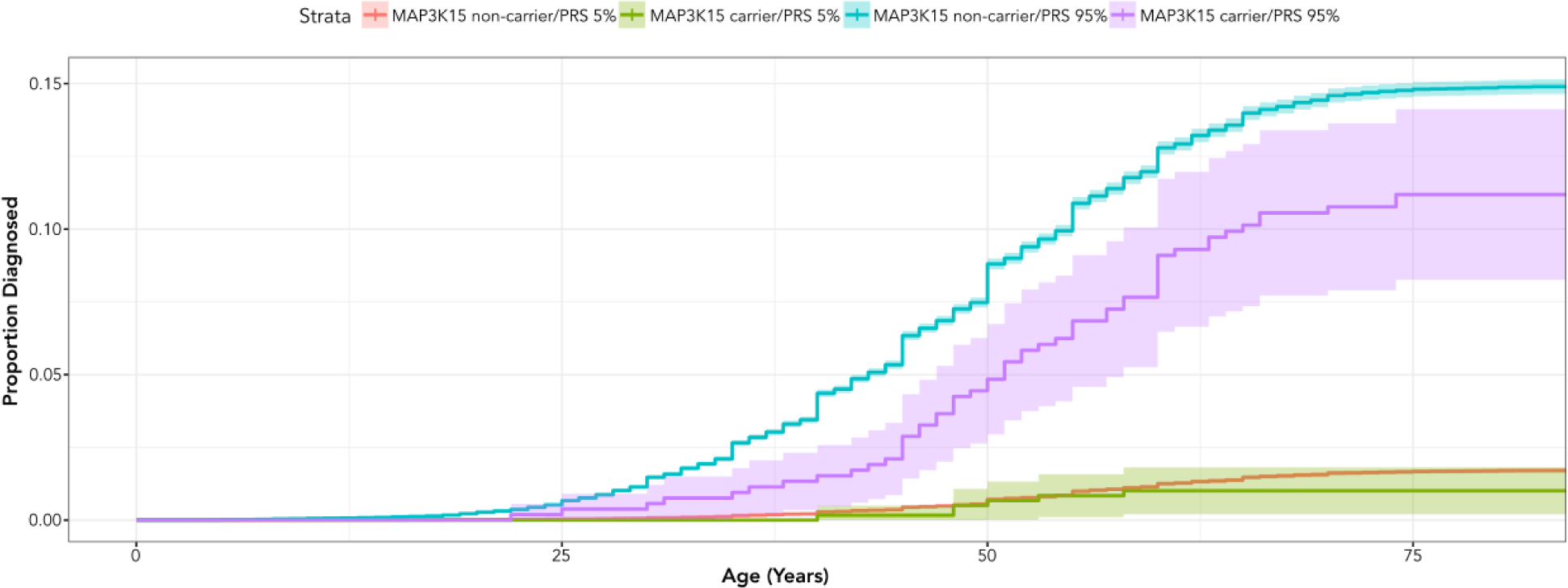
Impact of MAP3K15 LoFs and polygenic risk on T2D diagnosis. Individuals from the 23andMe database were stratified into high (top 5%) and low (bottom 5%) risk on the basis of their common variant derived T2D PRS. Within high and low risk PRS groups, individuals were further stratified by their imputed MAP3K15 carrier status. The cumulative incidence plot shows the cumulative proportion of individuals diagnosed with T2D by age, with the shaded region representing the 95% confidence intervals.

We were particularly interested in whether clinical lab values from 23andMe MAP3K15 LoF research participants would support the protective T2D and cholesterol associations. Obtaining a general overview of how genetic variants may influence metabolic health is achievable by profiling well-established and readily available blood biomarkers for metabolic disease. The relative ease of obtaining a blood metabolic panel enables the use of clinical laboratory data to validate genetic associations derived from self-reported phenotypic data, without the need to deploy custom devices or obtain access to highly specialized instrumentation. In order to assess the feasibility of using clinical laboratory values to replicate genetic associations and validate novel findings for MAP3K15, we developed a genetics-driven recruitment (GDR) capability from a small cohort among 23andMe research participants. Our goal was to assess whether we could 1) replicate the previously reported association of MAP3K15 LoF with decreased HbA1c and 2) validate the novel MAP3K15 LoF protective association with high cholesterol.

23andMe has conducted research studies in the past, where study participants were recruited on the basis of a condition they were already aware of or known genetic risk status disclosed to them in a 23andMe product report^2,33,34,35^. To further inform our understanding of the effect of MAP3K15 LoF on metabolic health, we established a protocol to recruit participants based on any type of genetics *unknown* to them, including the genotype(s) of a single variant or multiple variants, polygenic risk score, and others. We obtained Institutional Review Board (IRB) approval of the “DNA-Driven Discoveries Study” protocol which provides flexible coverage for multiple conditions and types of data collection and the return of individual-level clinical laboratory results to participants. As part of the study protocol, participants provide biological samples for clinical laboratory testing and biobanking, and answer survey questions related to medications, lifestyle, and disease history. Additionally, participants consent to sample biobanking for 10 years and give permission for 23andMe to share their de-identified data with qualified research collaborators. Participants may opt out at any time.

Our pilot study centered on the X chromosome MAP3K15 variant rs148312150, implicated in both LoF-burden testing as well as the 23andMe T2D GWAS. Of all the MAP3K15 LoF SNPs genotyped/imputed at 23andMe or genotyped in the UK Biobank, rs148312150 has the largest minor allele frequency (MAF = 0.453% in the 23andMe European cohort) and is the highest frequency MAP3K15 LoF protective association observed in burden tests for T2D and cholesterol. Examination of the 23andMe database showed over 4,000 consented hemizygous males and 58 consented homozygous females.

We worked with a third party provider to coordinate survey data collection, sample collection and processing, and return of laboratory results to study participants via an online portal. An overview of the recruitment and data collection process is shown in **Figure 4**. Participants who carry the MAP3K15 LoF variant were recruited first; age and sex-matched controls were then obtained via an identical process, resulting in a total recontacting of 5,600 individuals. We achieved a final study completion rate of 1.26%, with a total of 71 participants completing their blood draw. Relevant recruitment metrics are shown in **Table 1**.

**Figure 4.**
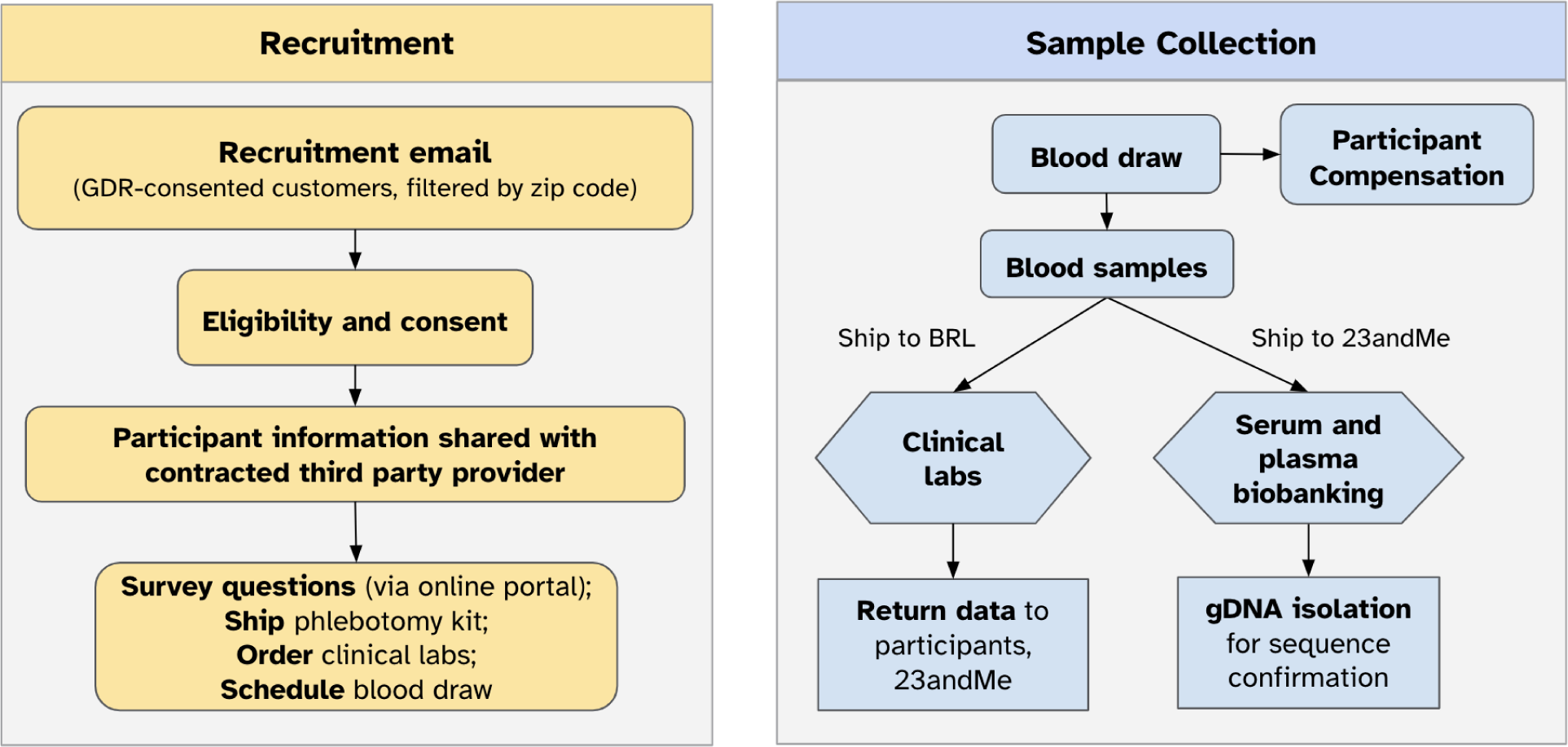
Recruitment and Sample Collection Workflow for the DNA-Driven Discoveries Study Pilot. *GDR* Genetics-Driven Recruitment, *BRL* BioReference Laboratories, *gDNA* genomic DNA

**Table 1:** The DNA-Driven Discoveries Study Recruitment Metrics for MAP3K15 LoF participants and age/sex-matched controls.

| # of People Contacted | Average Study Enrollment Rate | Average Study Completion Rate |
| --- | --- | --- |
| 5,600 | 1.87%<br>(106 enrollees) | 1.26%<br>(71 blood draws completed) |

While study enrollment was limited to residents within the zip-code range of the phlebotomy service, mobile phlebotomy enabled convenient study participation across the continental United States. The age demographics of our study participants tracked well with the distribution observed in the 23andMe database, with a median age of 48 years among all consented 23andMe research participants, and a median age of 52 years in our research study cohort (**Figure 5a, b**). The ancestry distribution of our participants also paralleled that of the 23andMe database (**Figure 5c**), with a slight enrichment of European individuals in our study population, potentially reflecting the higher minor allele frequency of rs148312150 in Europeans compared to other ancestries (MAF in Europeans: 0.453%; all other ancestries MAF < 0.05%).

**Figure 5.**
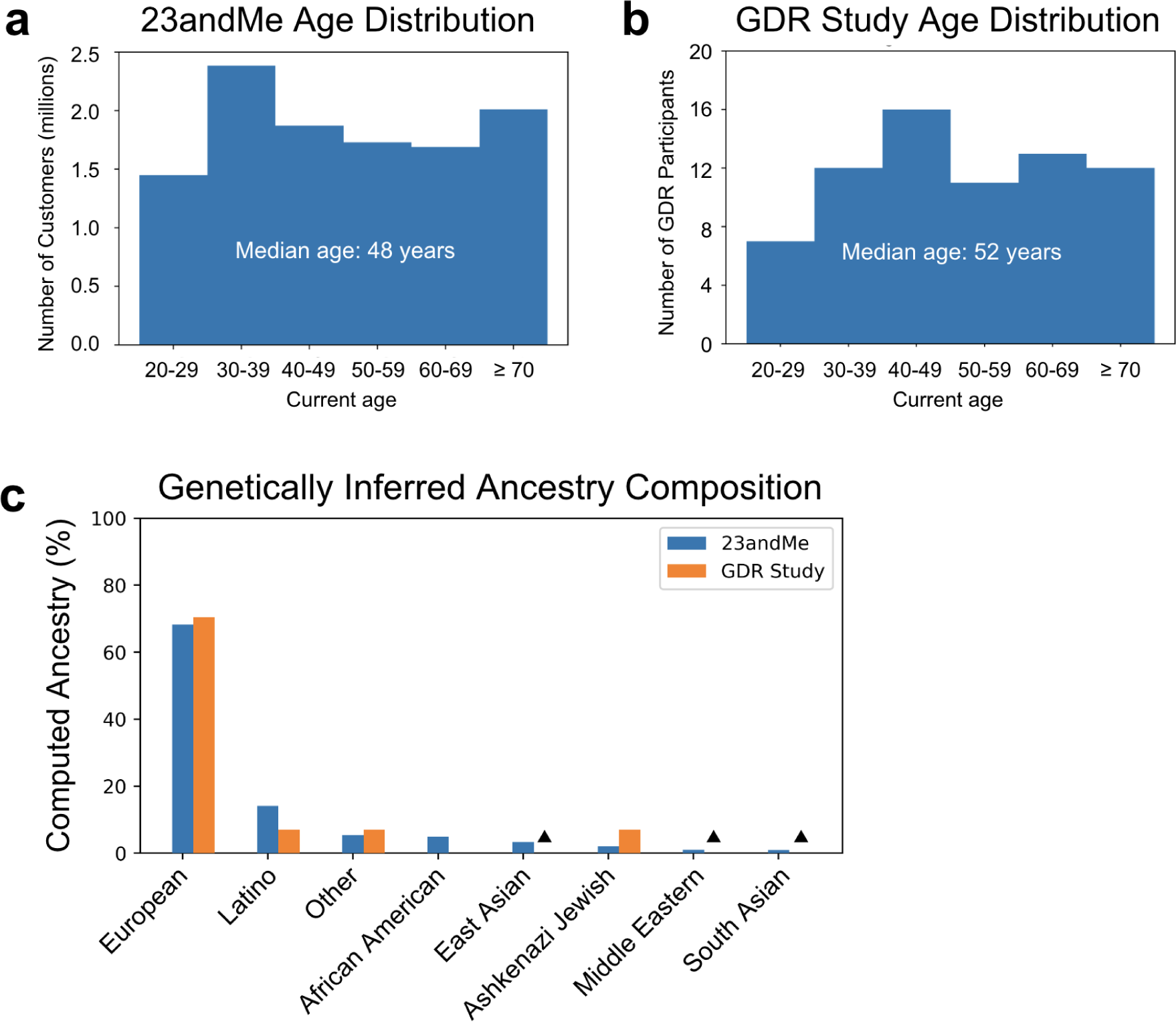
Age distribution and ancestry composition of DNA-Driven Discoveries Study participants compared to consented participants in the 23andMe database. (a) Age distribution of the research-consented 23andMe database (n ∼ 11 million). (**b)** Age distribution of pilot study participants who had completed a blood draw (n = 71). **(c)** Genetically inferred ancestry composition of the research-consented 23andMe database and pilot study research participants. ‘**▲**’ denotes categories with n<5 individuals, to protect participant privacy.

For the pilot cohort, we collected blood samples and obtained blood pressure (taken while the participant was seated, prior to their blood draw) along with a survey of personal/family medical history and participant lifestyle (diet, physical activity, medications). Blood samples were sent for clinical laboratory testing to obtain biomarker values relevant for metabolic health which included a lipid panel for various lipid species (e.g. cholesterol, LDL, triglycerides), glucose and HbA1c, kidney and liver panels, and a complete blood count. We also included clinical laboratory values for apolipoprotein B, high sensitivity C-reactive protein, and lipoprotein(a), which are highly relevant for cardiovascular health but not commonly included on standard wellness panels offered by a typical healthcare provider. The complete clinical laboratory panel assessed for each study participant is shown in **Extended Data Table 2**. To aid in the interpretation of laboratory results, participants were also asked whether they had taken medications prior to the blood draw and the identity of those medications.

While enrollment was open to male and female participants, we focused our control age-matched recruitment and subsequent data analysis on hemizygous males, owing to the lack of any homozygous female participant enrollment (likely attributable, in part, to low population frequency). Age-matching, where each knockout individual was age-matched within five years to a control, resulted in 31 participants per group successfully completing the blood draw and answering survey questions.

We examined our study cohort for evidence of clinical manifestation that could inform how MAP3K15 LoF contributes to protection against T2D, particularly in the form of the well-established T2D biomarker, HbA1c. We did not detect a statistically significant difference in HbA1c between MAP3K15 knockouts and controls (**Figure 6a**), possibly owing to the limited sample size of 31 individuals per cohort. By contrast, the UK Biobank data used to demonstrate a favorable trend for decreased HbA1c (effect: −0.085) had substantially more MAP3K15 LoF-carrying individuals: 1,900 homozygotes/hemizygotes, 5,185 heterozygotes, and 379,060 controls^25^. We observed a directionally-consistent effect size of 0.2 s.d. units, however, power calculations revealed that a minimum sample size of 350 individuals per group would be required to detect a statistically significant difference of this effect (i.e., 0.07% HbA1c). We further examined blood pressure data from our participants and observed a trend towards elevated median systolic blood pressure in MA3K15 LoF carriers compared to controls (130 mm Hg versus 120.5 mm Hg, p = 0.245), consistent with experimental observations in MAP3K15 knockout mice^29^, but in contrast to the analysis of UK Biobank data shown in Nag *et al.,* 2021^30^. There was no appreciable difference in median diastolic blood pressure in controls (79 mm Hg) compared to MAP3K15 LoF carriers (80.5 mm Hg).

**Figure 6.**
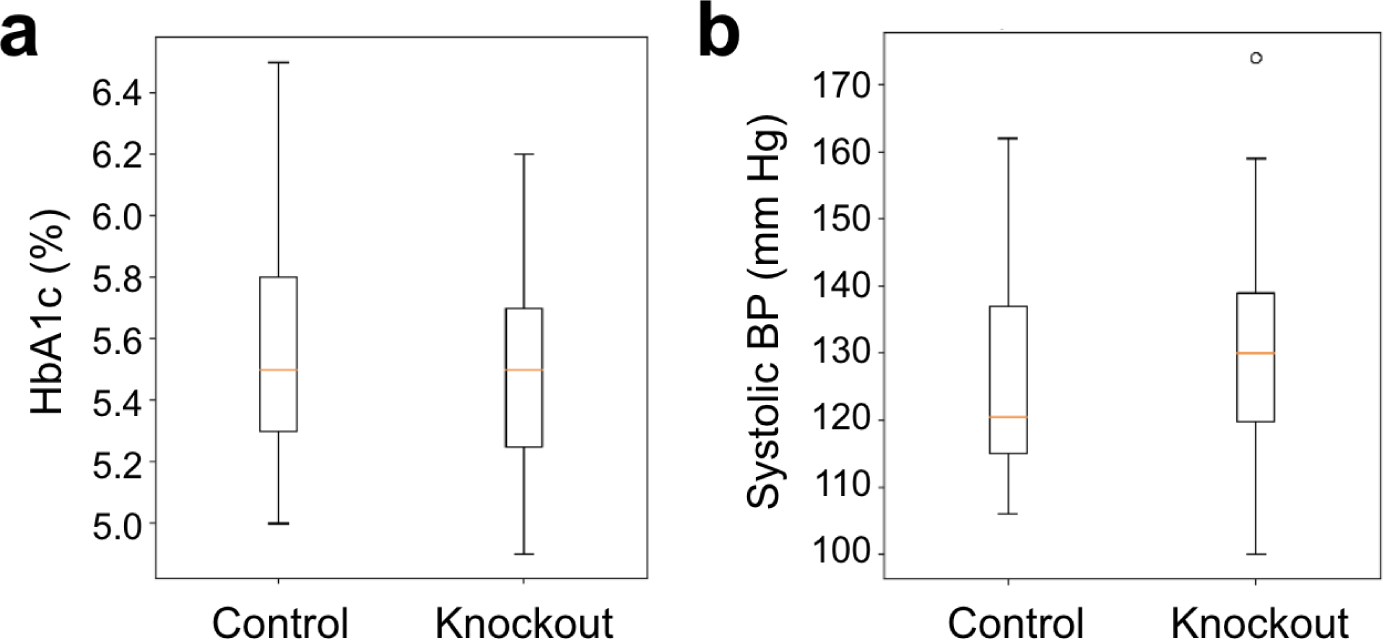
Summary of HbA1c and blood pressure values for DNA-Driven Discoveries Study research participants. (a) HbA1c, N = 31 per group. **(b)** Systolic blood pressure (BP), N = 28 per group. Data shown as median, interquartiles, and range.

We extended our investigation beyond the previously published associations with diabetes to determine whether lipid clinical laboratory values of our research participants supported the novel MAP3K15 LoF protective association with high cholesterol. A summary of relevant lipid panel data is shown in **Figure 7**. We observed marginal statistical significance for cholesterol (p = 0.0495, **Figure 7a**), consistent with the protective MAP3K15 LoF burden association with self-reported high cholesterol. We also observed a decrease in LDL/HDL ratio in MAP3K15 LoF carriers (p = 0.0444, **Figure 7f**). These data suggest that MAP3K15 LoF not only offers protection against total cholesterol levels, but also influences lipid composition to favor enrichment of HDL over LDL.

**Figure 7.**
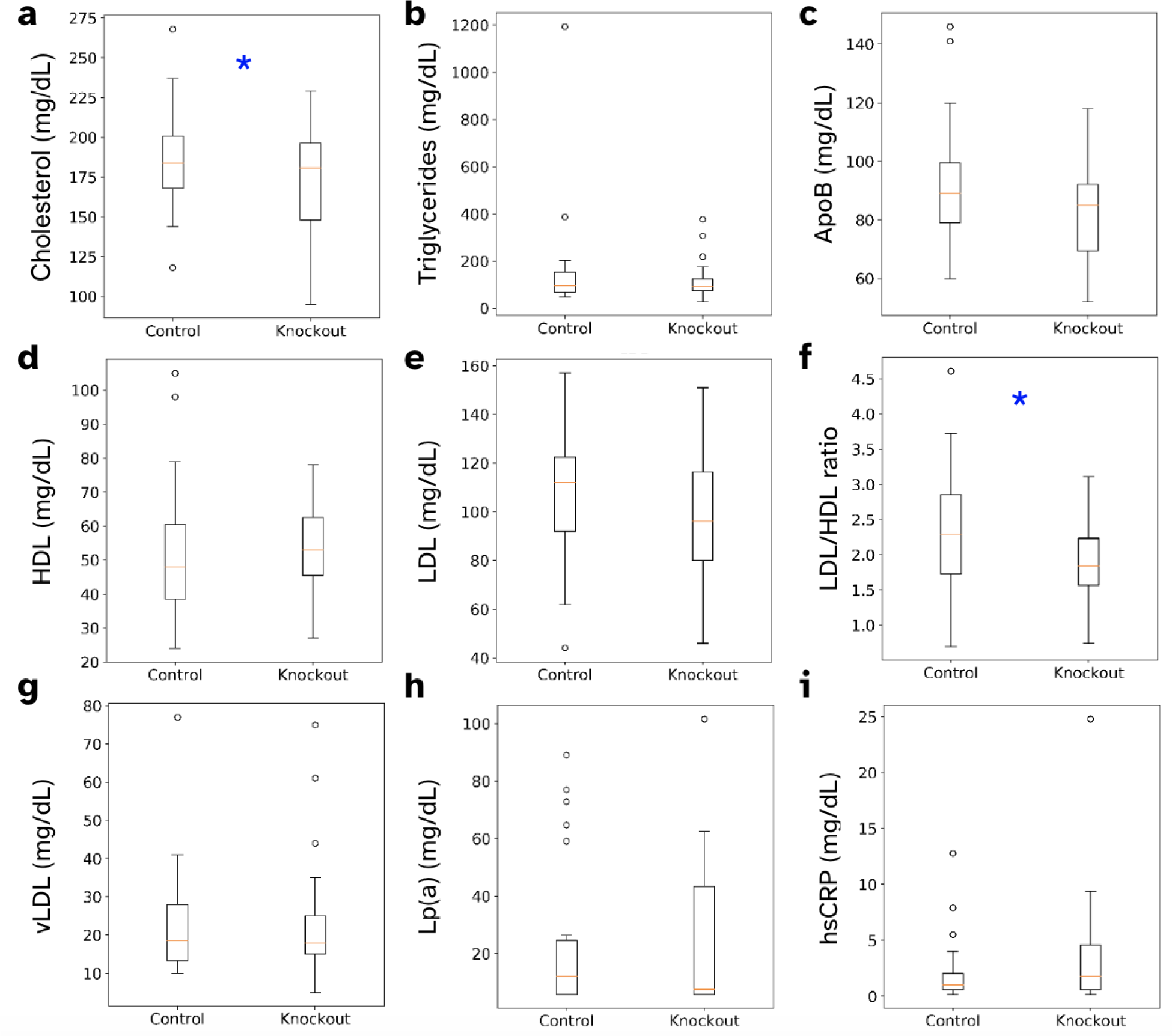
Lipid laboratory values comparing controls and MAP3K15 knockouts from DNA-Driven Discoveries Study research participants. *vLDL*: very low-density lipoprotein; *HDL*: high density lipoprotein, *ApoB*: apolipoprotein B; *Lp(a)*: lipoprotein(a). N = 31 per group; (* p < 0.05, Mann-Whitney U test, not adjusted for multiple testing, see Methods). Data shown as median, interquartiles, and range.

Despite the widespread availability of lipid laboratory values for UK Biobank research participants, studies using those data to correlate LoF variants and favorable effects on cholesterol laboratory phenotypes have failed to detect association with MAP3K15. We considered whether data from younger 23andMe participants could offer an explanation for the observed discrepancy, particularly in the age <40 group which is absent in the UK Biobank cohort. This is in part motivated by the observation that for many diseases, genetic relative risk declines with increasing age: in these situations genetic associations have stronger explanatory power (effect size) in younger age populations compared to older age populations^36^. We stratified each of our clinical laboratory results by age group and found a trend towards MAP3K15 LoF having a stronger protective effect against elevated cholesterol in participants aged 20 to 39 (**Figure 8a**). Among these individuals, the median cholesterol level was 189 mg/dL in control participants and 155 mg/dL in knockout participants (p = 0.065). We also observed a significant reduction in other LDL-related traits among younger individuals - among those aged 20-39, the median LDL/HDL ratio was 2.9 in control participants and 1.8 in knockout participants (**Figure 8b**, p = 0.04) and this difference was also seen among those in those in the age 40 to 59 age group (**Figure 8b**, p = 0.0087, with a median LDL/HDL ratio of 2.6 in control and 1.9 in knockout participants). The cardiovascular disease biomarker ApoB also showed trends of lower median values in MAP3K15 LoF carriers compared to controls: 101 mg/dL versus 84 mg/dL (p = 0.065) in participants aged 20 to 39, and 94 mg/dL versus 77.5 mg/dL (p = 0.090) in participants aged 40-59 (**Figure 8c**). The median laboratory value for these lipid traits among research participants aged 20 to 59 was consistently lower in MAP3K15 LoF carriers than in controls, suggesting that age effects are a potentially important consideration for these biomarkers in the context of protection associated with MAP3K15 LoF.

**Figure 8.**
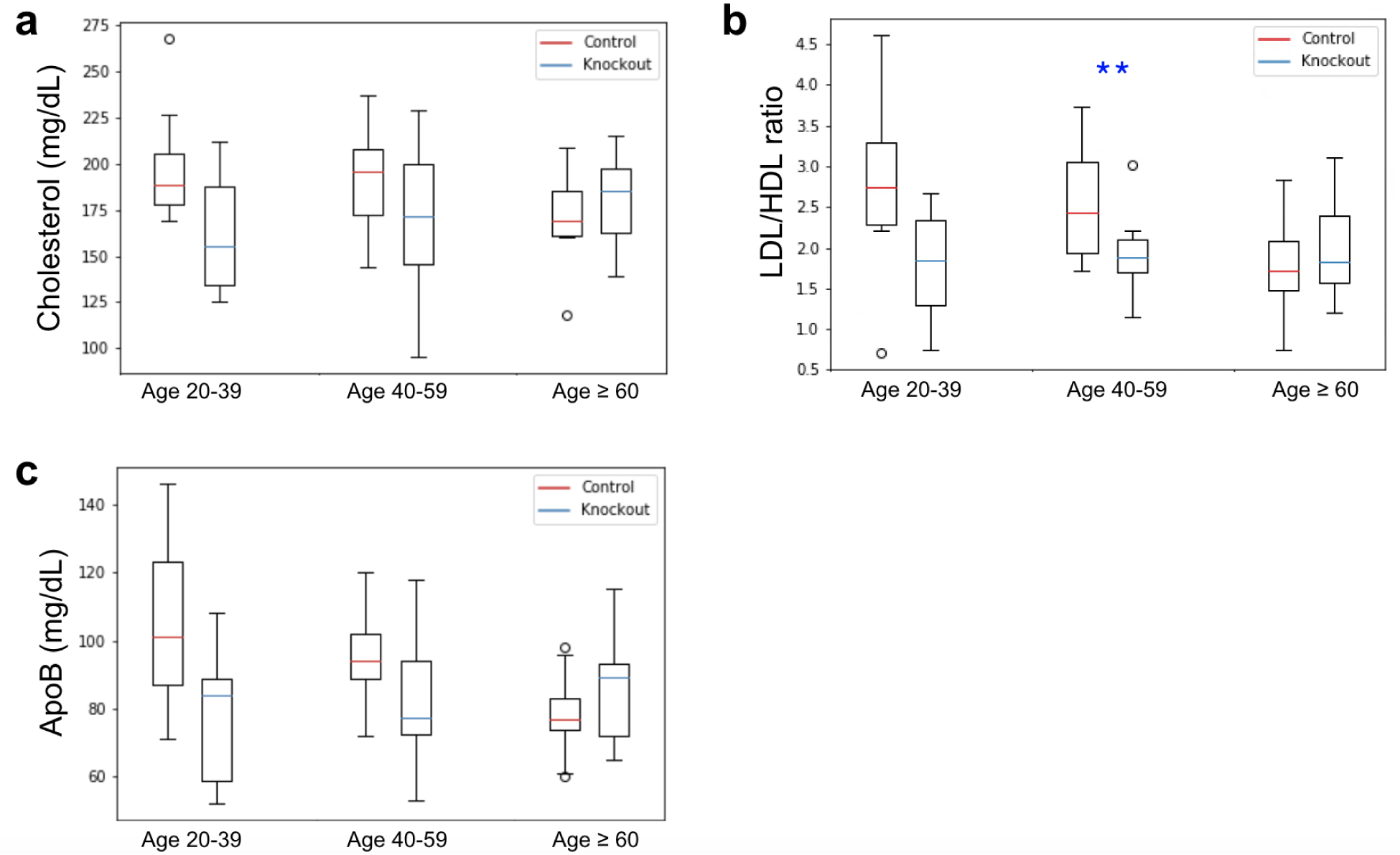
Age-stratified lipid laboratory values for (a) cholesterol, (b) LDL/HDL ratio and (c) apoB, comparing controls and MAP3K15 knockouts from DNA-Driven Discoveries Study research participants. Age 20-39 (n = 8), age 40-59 (n = 12), age ≥ 60 (n = 11); (** p < 0.01; Mann-Whitney U test). Data shown as median, interquartiles, and range.

## Discussion

Metabolic diseases such as T2D, obesity, and cardiovascular disease is a growing global concern. The prevalence of metabolic diseases has steadily increased over the past 20 years, alarmingly accompanied by strong sex, geographical, and socioeconomic disparities in mortality^37^. Leveraging genetics to discover and inform potential therapeutic targets such as MAP3K15 may provide additional information for consideration in drug development.

Our finding that MAP3K15 LoF is associated with a 4.5-year delay in the median age of onset of T2D among those at high polygenic risk to T2D is of potential clinical relevance for patients and of biological relevance for understanding disease susceptibility. Importantly, a 4.5-year delay in the onset of T2D diagnosis can make a difference in whether a patient ultimately goes onto develop a severe diabetic complication (e.g., diabetic retinopathy, diabetic nephropathy, or diabetic neuropathy) within their lifetime. It would be of considerable interest to monitor a small cohort of currently undiagnosed individuals with high versus low polygenic risk of T2D to see if and when (at what age) they are ultimately diagnosed with T2D. This would attest to the predictive value of MAP3K15 LoF (i.e., whether MAP3K15 LoF carriers can actually expect a delayed T2D diagnosis on average) to complement the suggested observation from our retrospective cohort data.

Based on MAP3K15 LoF burden analyses from the UK Biobank, Nag *et al.*^31^ did not identify any potential safety concerns associated with MAP3K15 inhibition. The 23andMe LoF burden results also do not indicate elevated blood pressure as a safety concern, however, MAP3K15 is implicated in opposite direction to T2D in the 23andMe data (**Figure 1**), and we observe a trend towards increased median systolic blood pressure readings in MAP3K15 LoF carriers from our GDR study cohort (**Figure 6b**). This demonstrates the complementary nature of analyzing both common and rare variants, and the relevance of deep phenotyping for validating genetic associations, all of which are well-enabled by the size of the 23andMe database.

The association of MAP3K15 LoF-mediated protection against high cholesterol should be considered hypothesis-generating and suggests potential additional therapeutic benefits (beyond diabetes) for targeting MAP3K15. While the effect size for protection against cholesterol is small, over time elevated cholesterol has a cumulative effect to impact cardiovascular disease, which is the leading cause of mortality worldwide and among individuals with type 2 diabetes^36^.

As a result of this study, we were able to establish and implement a successful recall framework to recruit consented 23andMe research participants on the basis of genetics unknown to them. This establishes a critical capability for 23andMe and collaborators for realizing the discovery potential of the 23andMe recontactable database.

Our pilot study had a final study completion rate of 1.26%. While study eligibility based on the geographic coverage of our mobile phlebotomy provider played a key role in the upfront attrition of our recruitable participant population, future studies could consider expanded phlebotomy coverage or partnering with local pharmacies for sample collection. IRB modifications leveraging compensation, recruitment methods (i.e., by postal mail instead of email), or more appropriately targeted recruitment or messaging may also improve enrollment. The ability to combine genetics-driven recruitment with other parameters from our deeply phenotyped 23andMe database allows focused and efficient recruitment on the basis of ancestry, comorbidities, geography, etc. An advantage of this combinatorial approach is that participant recruitment can be customized to improve diversity and representation in science.

When developing the DNA-Driven Discoveries Study protocol, it was important to us to enable the return of clinical laboratory results to our research participants. These clinical laboratory values revealed new clinically actionable findings for several phenotypes that participants did not self-report in their study survey responses. Blood pressure measurements revealed 32% of participants had undiagnosed Stage I or Stage II hypertension. Clinical laboratory values revealed 35% of participants were prediabetic (HbA1c ≥5.7% without self-reporting a diabetes diagnosis). A small fraction (n<5) of our pre-diabetic participants were under the age of 45, where laboratory results from this study may be their first indication of prediabetes. In the absence of other risk factors such as being overweight or having a family history of diabetes, most individuals will not be tested for HbA1c until age 35. Early information is beneficial for these individuals as prediabetes is reversible with lifestyle changes such as diet, increased activity, and weight reduction. The return of clinically actionable laboratory results may empower DNA-Driven Discoveries Study research participants to follow-up with their healthcare provider and inform their own healthcare decisions.

In addition to informing individuals of their health metrics, observations from the DNA-Driven Discoveries Study also highlighted important considerations for interpreting and analyzing metabolic GWAS. While discrepancies in power between the 23andMe database and the UK Biobank may explain our ability to detect a protective association of MAP3K15 LoF and high cholesterol, another intriguing difference between the two databases is the age distribution of their respective study participants. UK Biobank participants range from age 40 to 69, while the 23andMe database is a younger cohort: over a third of research participants are below the age of 40 (**Figure 5a**). Genetic analyses of younger participants may be subject to fewer confounders than in older populations, owing to variables such as age-gated prescription medication usage and shorter duration of exposure to environmental insult or lifestyle factors. Specifically, the interpretation of lipid laboratory data for individuals age greater than 40 may be confounded by statin usage and mask cholesterol protective phenotypes. Statins are commonly prescribed to diabetics above the age of 40 to decrease risk of cardiovascular events^38^. Patients with risk factors for cardiovascular disease are also recommended to start statins at a minimum age of 40^39^, and in the absence of other risk factors, statin therapy is typically not recommended for young adults^40^. Age-stratification or controlling for statin usage should be considered for future target discovery campaigns related to cholesterol phenotypes, as these adjustments may more accurately reflect genetic contribution to disease and improve discovery power.

An important conclusion from our study is that even small-scale studies can surface interesting observations, nominating new testable hypotheses or ways to refine phenotype definitions or GWAS analysis to inform drug development and genetic research. Indeed, observations from our pilot study prompted us to dive more deeply into the database for insight into MAP3K15 LoF-mediated protection against high cholesterol and T2D, with implications for how we interpret and analyze self-reported high cholesterol phenotypes (e.g., age-stratification of GWAS or LoF burden testing for target discovery) or conduct future data collection (e.g., requesting non-diabetic research participant HbA1c values and monitoring time to T2D diagnosis from currently non-diabetic individuals).

Genetics-driven recruitment studies select participants based on specific genetic profiles, in contrast to GWAS, which focus on phenotypes. However, GDR studies require careful consideration of multifaceted and significant ethical concerns, primarily centered on balancing participants’ autonomy and their “right not to know”^41^. We evaluated the pros and cons of sharing information about the participant’s role as a case or control, individual-level clinical laboratory results, and exploratory genetic data with the participants. We assessed the risks, including informational harm, emotional discomfort without actionable results, and risk associated with providing clinically unvalidated information. After careful consideration, and determining that the benefit of sharing information did not outweigh the risks, we decided not to inform participants of their case or control status due to the exploratory nature of the research. However, we offered the return of clinical laboratory values, which participants could choose to access and follow up on with a healthcare provider, in case they might be actionable results. We implemented these safeguards to ensure participants’ well-being and interpretation of results within a trusted patient-physician relationship.

Future studies using the DNA-Driven Discoveries Study protocol can enable similar recruitment and workflows, customized for a variety of disease areas, data/sample collection, and research applications, including recruitment of family members to increase power for especially rare variants (cascade screening), or attesting to the validity and interpretation of self-reported data in the 23andMe database via correlation with clinical laboratory values or relevant biomarkers. In addition, the collection of laboratory values and biobanked samples from the 23andMe Research Cohort may serve as a useful internal validation dataset. These data may be more reflective of the United States population than other repositories such as the UK Biobank and FinnGen. It is our hope that expanded use of the DNA-Driven Discoveries Study capability will improve the success of drug development for patients and give back to the research participants, for which the value of their altruism, trust, and willingness to participate in genetic-driven recruitment studies cannot be overstated.

## Acknowledgments

We are grateful to the 23andMe customers who consented to participate in research for enabling this study. We also thank Bill Richards and Adam Auton as members of the 23andMe Therapeutics Leadership Team for their support and feedback, and 23andMe employees who contributed to the development of the infrastructure that made this research possible, particularly Sarah Elson for conceptualizing and enabling GDR opt-in in 2018, and individuals who participated in our internal employee pilot.

## Author contributions

J.J.B. conceptualized the study. J.J.B., K.K., and D.N. designed and implemented the study. K.K. and S.D. coordinated recruitment and sample/survey data collection with our third party provider. D.N. wrote the study protocol. J.J.B. and Y.Q. collected data and processed samples. J.J.B., J.W., and Z.L.F. analyzed data, in consultation with X.W.. J.J.B., X.W., Z.L.F., and D.N. wrote the manuscript with input from M.V.H., L.S., and S.S..

The following members of the 23andMe Research Team contributed to this study: Stella Aslibekyan, Adam Auton, Elizabeth Babalola, Robert K. Bell, Jessica Bielenberg,

Jonathan Bowes, Katarzyna Bryc, Ninad S. Chaudhary, Daniella Coker, Sayantan Das, Emily DelloRusso, Sarah L. Elson, Nicholas Eriksson, Teresa Filshtein, Pierre Fontanillas, Will Freyman, Zach Fuller, Chris German, Julie M. Granka, Karl Heilbron, Alejandro Hernandez, Barry Hicks, David A. Hinds, Ethan M. Jewett, Yunxuan Jiang, Katelyn Kukar, Alan Kwong, Yanyu Liang, Keng-Han Lin, Bianca A. Llamas, Matthew H. McIntyre, Steven J. Micheletti, Meghan E. Moreno, Priyanka Nandakumar, Dominique T. Nguyen, Jared O’Connell, Aaron A. Petrakovitz, G. David Poznik, Alexandra Reynoso, Shubham Saini, Morgan Schumacher, Leah Selcer, Anjali J. Shastri, Janie F. Shelton, Jingchunzi Shi, Suyash Shringarpure, Qiaojuan Jane Su, Susana A. Tat, Vinh Tran, Joyce Y. Tung, Xin Wang, Wei Wang, Catherine H. Weldon, Peter Wilton, Corinna D. Wong.

## Data availability statement

Given restrictions to the availability of 23andMe individual-level data owing to 23andMe consent and privacy/IRB guidelines, data sharing is limited to aggregate statistics that are necessary for reproducibility. Broader data sharing pertaining to the data/samples supporting the findings of this study may be made available upon reasonable request or collaboration, to qualified researchers under an agreement with 23andMe that protects the privacy of the 23andMe participants, in accordance with individual consent and additional requirements within the scope of our IRB approval.

## Extended Data

**Extended Data Table 1.** MAP3K15 LoF burden test associations and relevant statistics from the 23andMe database. *OR*: Odds ratio, *CI*: Confidence interval

| Phenotype | p-value | OR | 95% CI |
| --- | --- | --- | --- |
| Type 2 diabetes | 3.6e-63 | 0.836 | 0.819-0.854 |
| Gestational diabetes | 7.5e-38 | 0.752 | 0.721-0.785 |
| Hyperglycemia | 3.0e-23 | 0.853 | 0.827-0.880 |
| High cholesterol | 2.2e-8 | 0.961 | 0.948-0.975 |
| Diabetic neuropathy | 2.4e-7 | 0.877 | 0.835-0.922 |
| Diabetic retinopathy | 5.6e-7 | 0.807 | 0.742-0.877 |

**Extended Data Table 2.** Summary of Clinical Laboratory Values Obtained from DNA-Driven Discoveries Study Research Participants.

| Liver/Kidney Panel | Whole Blood Cell Count | Glucose |
| --- | --- | --- |
| Total Protein<br>Albumin<br>Globulin<br>Albumin/Globulin Ratio<br>Sodium<br>Potassium<br>Chloride<br>Carbon Dioxide<br>Blood Urea Nitrogen<br>Creatinine<br>Estimated Glomerular Filtration Rate<br>Calcium<br>Bilirubin<br>Lactate Dehydrogenase<br>Alkaline Phosphatase<br>Alanine Transaminase<br>Aspartate Transaminase<br>Gamma-glutamyl Transpeptidase | White Blood Cells<br>Red Blood Cells<br>Hemoglobin<br>Hematocrit<br>Mean Corpuscular Volume<br>Mean Corpuscular Hemoglobin<br>Mean Corpuscular Hemoglobin Concentration<br>Red Blood Cell Distribution Width<br>Polymorphonuclear Cells (Neutrophils)<br>Lymphocytes<br>Monocytes<br>Eosinophils<br>Basophils<br>Immature Granulocytes<br>Platelet Count<br>Mean Platelet Volume | Glucose<br>Hemoglobin A1C |
|  |  | Lipid Panel |
|  |  | Cholesterol<br>Triglyceride<br>High-Density Lipoprotein<br>Low-Density Lipoprotein<br>Very-Low-Density Lipoprotein<br>Lipoprotein (a)<br>Apolipoprotein B |
|  |  | Cardiovascular |
|  |  | Blood Pressure (measured)<br>C-Reactive Protein (high sensitivity) |

